# TRiXi: A Multi-Target Tandem Repeat-Based Method for Accurate Detection of X-Inactivation Skewing in Humans

**DOI:** 10.1101/2025.11.24.25340860

**Authors:** Dóra Goldmann, Björn Gylemo, Maike Bensberg, Ingela Johansson, Svenja Löffert, Júlia Goldmann, Lisa Haglund, Magdalena Vitkova, Janos Kondri, Sandra Hellberg, Engla Haglund, Shadi Jafari, Johnny Ludvigsson, Colm E. Nestor

## Abstract

Preferential usage of either X-chromosome, skewed X-inactivation (sXCI), can profoundly alter the penetrance and expressivity of X-linked traits in females, often with life-threatening consequences. Despite its clinical relevance, the true frequency, origins, and stability of sXCI in human tissues remain poorly understood, in part due to a lack of effective and sensitive methods for assessing XCI status. Here, we present TRiXi (<u>T</u>andem <u>R</u>epeat-based Identification of <u>X</u>-Inactivation), a simple and highly sensitive technique capable of detecting sXCI from as little as 10 ng of archived human DNA. Applying TRiXi to over 1,000 neonatal cord blood samples from healthy infants, we demonstrate that extreme sXCI is a common constitutional feature in humans. Use of longitudinal samples from the same donors revealed that sXCI patterns in skewed neonates were stably maintained into adolescence, whereas in non-skewed females, patterns fluctuated considerably over time. Using low-pass short-read sequencing, whole-genome long-read sequencing, and targeted exon sequencing, we exclude most known genetic drivers of sXCI in extremely skewed infants, supporting stochastic processes as the primary origin of skewed XCI. Further, by inferring X-inactivation patterns in 4,571 adult tissues from 264 females in the GTEx cohort, we demonstrate that skewed XCI is not restricted to blood, revealing widespread multi-organ sXCI in females. Our study represents the most comprehensive analysis of constitutional sXCI to date and reveals sXCI as a widespread and stable epigenetic trait. This can profoundly modify the penetrance of X-linked conditions with immediate implications for genetic diagnostics and our understanding of human disease variability. TRiXi provides a convenient and accessible approach to inform diagnosis of X-linked traits by assaying sXCI. Studying individuals with sXCI may yield critical insights into (1) the initiation of XCI and its underlying genomic elements, (2) previously unrecognized lethal X-linked traits and (3) improved polygenic risk prediction through incorporation of X-chromosome variation.

## INTRODUCTION

Typically, human females possess two X-chromosomes (46, XX) whereas males have one X-chromosome and one Y-chromosome (46, XY). To prevent lethality due to a double dose of X-linked genes in females, one X-chromosome is inactivated in all female cells during the peri-implantation stage of human development^1^. X-chromosome inactivation (XCI) is initiated by expression of the long non-coding RNA, *XIST*, from one X-chromosome. *XIST* progressively coats the chromosome from which it is expressed, which in turn becomes marked with DNA methylation and repressive histone marks, ultimately resulting in compaction and transcriptional silencing^2–4^. The choice of parental X-chromosome to undergo XCI in each cell is random; different cells in the same female will have inactivated the paternal X-chromosome (Xp) or maternal X-chromosome (Xm). Although the initial choice of X-chromosome for inactivation is random in humans, the inactivated X-chromosome is stably inherited in a clonal fashion throughout all subsequent cell divisions^5^. Consequently, women are ‘mosaic’ for XCI, typically exhibiting a ratio of ∼1:1 between cells having inactivated Xp or Xm^6^.

The mosaic nature of XCI generally reduces the penetrance and expressivity of traits encoded on the X-chromosome in females, as roughly half of their cells express the wild-type allele, which compensates for the effects of the mutant allele in the remaining cells. In contrast, many X-linked diseases, such as Rett syndrome and Fragile X syndrome, are considerably more severe and often embryonic lethal in males due to mono-allelic expression of the mutated X-linked gene^7^. However, females can manifest similarly severe and life-threatening symptoms of X-linked disorders due to skewed X-inactivation (sXCI), a state in which most, or all, cells within or across tissues have inactivated the same parental X-chromosome. While much of the sXCI observed in adult females results from secondary selection in the form of clonal hematopoiesis, which tends to increase with age, we and others have reported that extreme skewing of XCI may also be a common and potentially constitutional genetic characteristic in humans^6, 8–12^. Clinically, constitutional sXCI would have a more profound and earlier effect on the penetrance and expressivity of X-linked traits in females. The determination of sXCI in bulk tissues such as blood is complex, requiring both an informative (heterozygous) genetic variant and an additional layer of information such as DNA methylation or gene expression. The most commonly used method for detection of skewing, the HUMARA assay, lacks sensitivity, failing to provide a result in 20-30% of samples tested^13^. The lack of an effective and sensitive approach to assay sXCI has limited our ability to characterize the prevalence and origin of XCI skewing in humans, a phenomenon predominantly studied in adult females, where constitutional skewing is often obscured by clonal cell expansion.

We introduce <u>T</u>andem <u>R</u>epeat-based Identification of <u>X</u>-chromosome Inactivation (TRiXi), an improved method for high-sensitivity detection of skewed XCI. Applying TRiXi to archived DNA from over 1,000 cord blood samples revealed that constitutional sXCI is surprisingly frequent in humans (>5%). Using a range of whole genome and targeted approaches we further investigate the genetic origin and stability of sXCI during childhood. Finally, through multi-omic analysis of ∼4,500 tissues from the GTEx project, we characterize sXCI patterns across healthy human adult tissues and reveal skewing as a powerful and far more frequent modifier of X-linked traits than previously appreciated.

## RESULTS

### TRiXi - an efficient method for high-sensitivity detection of skewed X-inactivation

Measuring the degree of sXCI in females requires the use of techniques that combine an informative genetic variant with an additional layer of molecular information such as DNA methylation or gene expression. Whereas a plethora of methods for sXCI detection have been proposed over the last decade^14–20^, the HUMARA assay, developed by Allen et al in 1992^21^, remains the gold-standard method in the field. The HUMARA assay involves methyl-sensitive restriction digest (only the unmethylated active X is cut) of the highly polymorphic short tandem repeat (STR) in the androgen receptor gene (*AR*; formerly HUMARA) followed by PCR amplification and electrophoresis to reveal XCI skewing; only the methylated, uncut inactive X allele will be amplified after digestion and the ratio between each allele represents XCI skewing in a given sample (**Supplementary Fig. 1a**). However, the HUMARA assay is uninformative in the 20-30% of females that are homozygous at the *AR* repeat. Moreover, the accuracy of the HUMARA assay is further affected by locus-specific alterations in DNA methylation, the presence of genetic variants in the target sequence, and preferential PCR amplification of the shorter allele^22^.

To overcome the limitations of the HUMARA assay, we developed TRiXi to enhance the detection of sXCI. Using a similar concept to the HUMARA assay we aimed to identify multiple polymorphic tandem repeats across both arms of the X-chromosome and combined these into a single multiplex PCR reaction, thereby increasing assay sensitivity while requiring no additional experimental steps, expense or equipment. Using whole genome sequencing (whole blood) from 50 randomly selected females from the GTEx project and whole genome methylation data (isolated T-cells) from four females in the BLUEPRINT project we identified all X-linked repeat sequences that (i) are highly polymorphic tri- and tetra-nucleotide repeats, (ii) are flanked by at least one *Hpa*II site, and (iii) are characterized by 50% methylation, indicating that the locus is subject to XCI (**Fig. 1a, b**, **Supplementary Table 1**). The *AR* locus was intentionally omitted from the TRiXi assay, as hyper-expanded *AR* repeats are common in the general population. Such long expansions are typically undetectable in the HUMARA assay, as they fall outside the expected size range and PCR amplification tends to favor the shorter, non-expanded (healthy) allele. The normal repeat length range of the *AR* locus is unusually broad (7–27 CAG repeats), often resulting in substantial allele length differences within individuals (**Fig. 1b**), increasing the probability of PCR bias due to size-dependent amplification efficiency^13^. In total, we identified six tri-and nine tetra-nucleotide repeats that satisfied all selection criteria (**Supplementary Table 1**). Through exhaustive iterative testing we succeeded in multiplexing five informative loci, allowing robust detection of all ten alleles in a single assay (**Fig. 1c, d, Supplementary Table 2**). We next performed both the HUMARA assay and TRiXi on 50 umbilical cord samples from the All-Babies in Southeast Sweden (ABIS) birth cohort^23, 24^. Results from TRiXi and HUMARA were highly correlated (**Fig. 1e, Supplementary Table 3**), but as expected, HUMARA failed to determine sXCI in 12 samples (12/50, 24%) due to homozygosity at the *AR* locus while TRiXi was informative in all samples (**Fig. 1f**). To further validate TRiXi we performed whole genome Oxford Nanopore Technologies (ONT) long-read sequencing on one highly skewed sample (97% sXCI; blue dot in **Fig. 1e, f**). Following phasing of reads based on repeat length, ONT long-read sequencing confirmed the extreme skewing observed in this sample (**Fig. 1g**). Further, using a recently published method for calculating skewing from X-chromosome ONT data^17^, resulted in a global X-chromosome skewing of 88.1%, again highly concordant with that obtained with TRiXi (97%).

**Figure 1.**
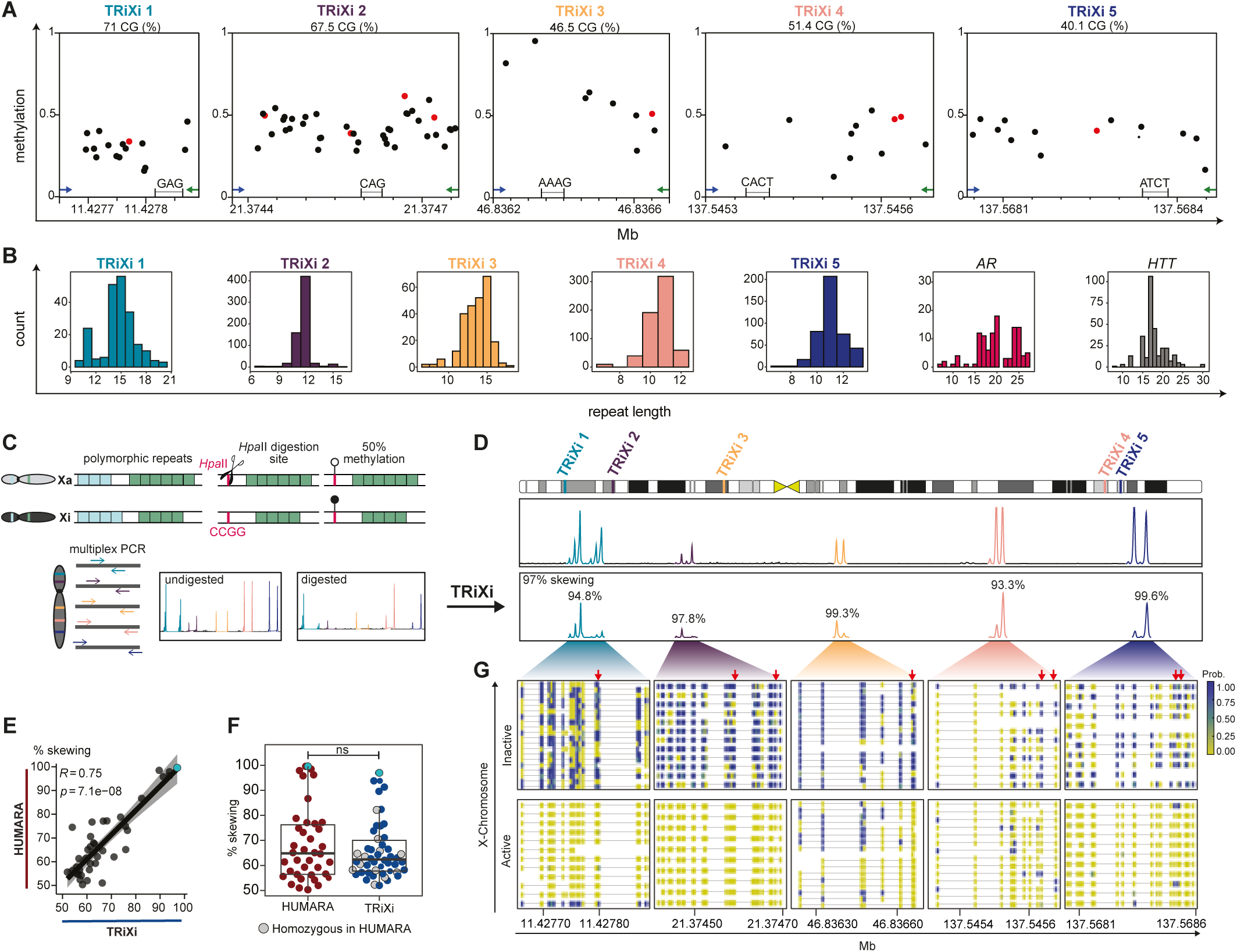
Development of <u>T</u>andem <u>R</u>epeat-based Identification of <u>X</u>-chromosome Inactivation (TRiXi) for high-sensitivity detection of skewed X-inactivation. a, forward (blue) and reverse (green) primer binding sites, repeat location (hg38), repeat sequence, PCR product GC content (%) and methylation of CpG sites in TRiXi 1-5 PCR target regions. *Hpa*II (CCGG) sites are highlighted in red. **b**, allele length distribution of all five TRiXi repeats estimated from 50 randomly selected female WGS samples from the GTEx dataset. The highly polymorphic *AR* (X-Linked) and *HTT* (autosomal) tandem repeats are shown for comparison. **c**, schematic illustration of the TRiXi method involving methylation-sensitive digest of the alleles on the active X-chromosome followed by multiplex PCR of all 10 alleles (five polymorphic loci) followed by fragment size determination by capillary electrophoresis. **d**, Representative results of a single TRiXi assay. Top and bottom panels show undigested and digested runs of a highly skewed female sample from the ABIS cohort (sample ABIS 1). **e**, Scatterplot of XCI skew of 50 neonatal samples determined by HUMARA and TRiXi. Spearman’s rank correlation test; ρ = 0.75, p = 7.1×10^−8^. **f**, Boxplot of XCI skew of 50 neonatal samples determined by HUMARA and TRiXi. Samples which were uninformative in HUMARA assay are indicated in grey. Two-tailed Wilcoxon rank-sum test; p = 0.68. **e**, **f**, light-blue dots highlight sample ABIS 1. **g**, Clustering of ONT reads by repeat length at all five repeat loci in the ABIS 1 sample. Color gradient indicates CpG DNA methylation level. Red arrows indicate CCGG sites.

### Over 5% of newborns exhibit extreme XCI skewing

We next set out to determine the frequency of constitutional skewing in humans by performing TRiXi on 500 archived umbilical cord blood samples from healthy infants in the ABIS birth cohort^23, 24^. We determined the true heterozygosity and allele frequencies for all five polymorphic loci, resulting in a TRiXi sensitivity of 99.82%—equivalent to only 1 in 500 females being homozygous at all five repeats (**Supplementary Fig. 1b**). Skewing estimates were highly concordant across all loci within each sample, supporting the use of DNA methylation patterns across both arms of the X-chromosome as reliable proxies for XCI status (**Supplementary Fig. 1c, Supplementary Table 4**). Whereas including five or more polymorphic repeats in the assay provides the highest sensitivity, fewer repeats can be used. Indeed, estimating sXCI in an additional 683 umbilical cord blood samples incorporating only the two most polymorphic repeats TRiXi 1 and TRiXi 2 offered a detection rate of 97% (21/683 samples homozygous for both repeats), a vast improvement over the HUMARA assay (**Supplementary Table 5**). Altogether, analyzing skewed X-inactivation in 1,027 neonates revealed that 10.42% of females were highly skewed (mean +1 SD, >71.09% skewing) for XCI at birth and 5.16% exhibited extreme skewing (mean +2 SD, >79.76% skewing) (**Fig. 2a**). To investigate whether repeat length affects the accuracy of XCI skewing measurement by TRiXi, we compared the distributions of the repeat lengths between mosaic, skewed and extremely skewed females, which showed very similar distributions (**Supplementary Fig. 1d**). Furthermore, similar skewing values were obtained for each individual repeat, indicating that, in the case of homozygosity of one or more repeats, TRiXi reliably reveals XCI skew (**Supplementary Fig. 1e**).

**Figure 2.**
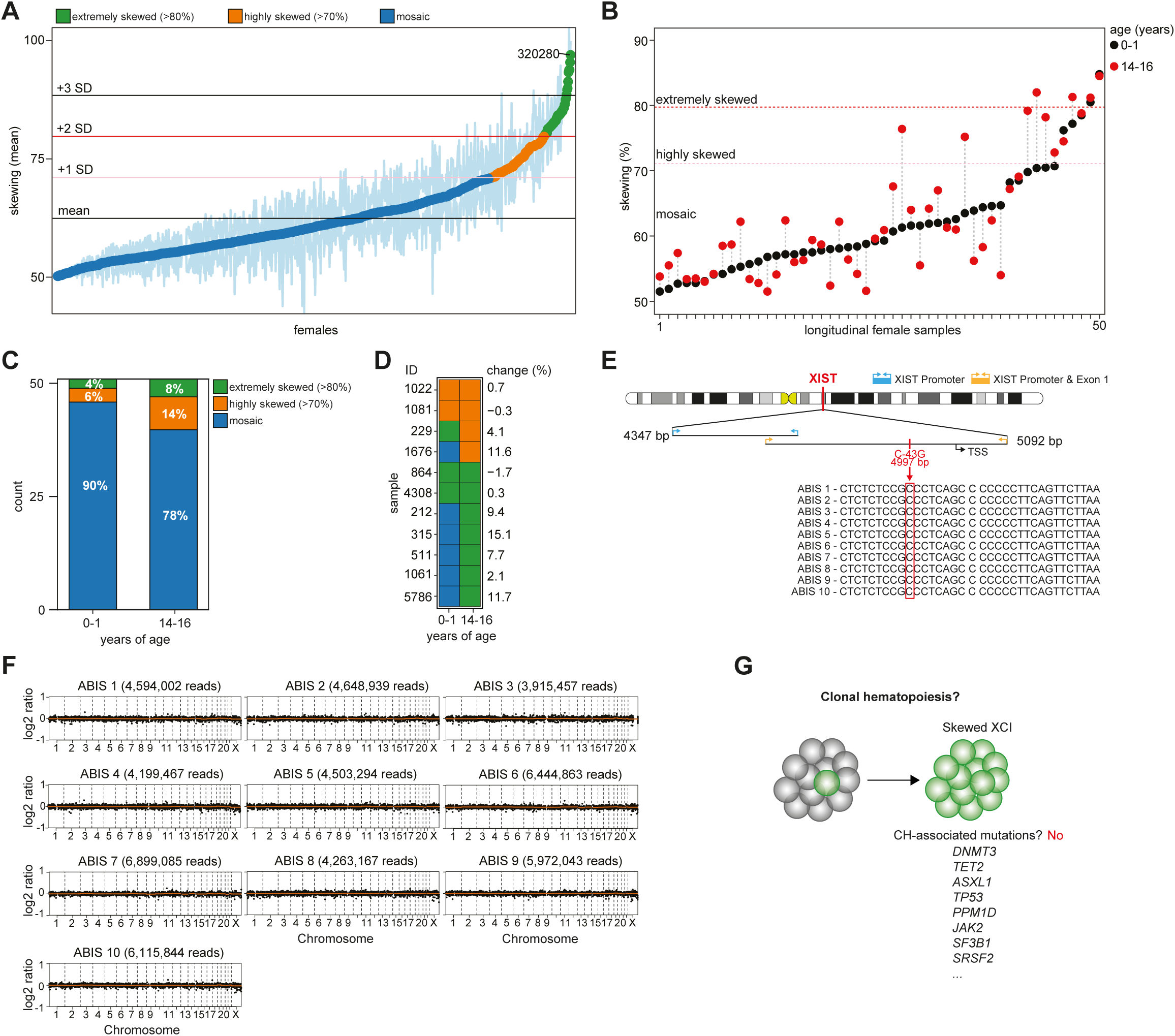
The distribution of skewed X-inactivation in 1,000 newborns revealed by TRiXi. a, XCI status of 1,027 neonatal blood samples determined by TRiXi. **b,** XCI skew of longitudinal samples collected between 0-1 years of age and between 14-16 years of age. **c**, Frequency of mosaic, highly or extremely skewed XCI at 0-1 and 14-16 years of age. **d**, classification and change (%) in skewing between timepoints for samples that were highly or extremely skewed at 14-16 years of age. **e**, *XIST* promoter region and first exon of 7 skewed samples (ABIS 1-7) and 3 controls (ABIS 8-10) investigated with sanger sequencing. Sequences spanning a previously reported variant (C-43G) associated with constitutional skewing highlighted in red. **f,** copy number analysis using low-pass whole genome sequencing data for the same skewed females. **g,** targeted exon sequencing of clonal hematopoiesis-associated genes revealed no mutations in any of the genes included in the oncoReveal™ Myeloid Panel.

To determine whether the observed skewing represented a stable, intrinsic property of the hematopoietic lineage or a transient phenomenon, we analyzed follow-up samples from 50 individuals at 14–16 years of age (**Fig. 2b, Supplementary Table 6**). The majority of females were classified as mosaic at both timepoints as 39 out of 50 (78%) females were mosaic at both at 0-1 and 14-16 years of age. Interestingly, the degree of XCI skew changed considerably (≤5%) in 16 out of 50 (32%) females, leading to an almost doubling of extremely and highly skewed females (**Fig. 2c**). Further, females that were classified as extremely skewed or skewed at age 0-1 exhibited relatively small changes in XCI skew, maintaining their skewed status into adolescence (**Fig. 2d**). These results suggest that in up to 30% of females, considerable selection of X-allele usage is occurring from birth.

### No clear genetic basis for skewed X-inactivation in neonates

The observation of pronounced skewing at birth, that is subsequently maintained throughout childhood suggests a primary or constitutional etiology. Organism-wide, extreme skewing may result from constitutional genetic variants that directly bias the inactivation process toward a specific parental X-chromosome. Examples include mutations in the *XIST* gene promoter or balanced X:autosome translocations. Similarly, disruptions of the X-inactivation center (XIC) or autosomal elements involved in X-chromosome choice have been implicated in promoting or directly mediating primary skewing of X-inactivation^25–27^. Using seven of the most skewed samples (ABIS 1-7) and three mosaic females (ABIS 8-10), we conducted a thorough investigation of the genetic origin of skewed XCI. We first performed tiled sanger sequencing across the *XIST* promoter but failed to identify previously reported or novel variants that could affect *XIST* expression (**Fig. 2e**). Similarly, low-pass short-read sequencing failed to identify copy number variations or aneuploidies recently reported in completely skewed females^6^ (**Fig. 2f**). The levels of skewing were highly variable during childhood (**Fig. 2b, c, d**), which could indicate an ongoing process of clonal selection that drive XCI skewing. Whereas clonal hematopoiesis (CH) is primarily studied in older individuals, mutations in key CH genes such as *TP53*, *JAK2*, *TET2* and *DNMT3A* are often observed in childhood malignancies and familial disorders^26, 28–30^. We thus investigated whether the observed extreme skewing could have resulted from intense CH due to a constitutional mutation in CH genes. Again, exon sequencing of the 58 most well-characterized CH genes failed to identify any previously identified CH-associated mutations in these genes (**Fig. 2g, Supplementary Table 7**). Finally, deep (40X) ONT long-read sequencing of the most skewed sample (**Fig. 1g**), failed to identify any clearly pathogenic X-linked or autosomal mutations that could explain the observed skewing, including an absence of balanced X:autosome translocations. Together, our genetic analysis of skewed females did not identify a definitive underlying cause, suggesting that the observed extreme skewing arose from purely stochastic processes or as of yet unknown genetic modifiers of XCI.

### Evidence of multi-organ constitutional sXCI in adult females

A recent cross-tissue analysis of XCI skewing suggested that XCI initiation occurs before germ-layer specification^31^. Whereas stochastic selection of a limited number of progenitor cells for lineage specification can lead to sXCI within a given tissue, it is highly unlikely that such a process would independently give rise to similar skewing across multiple, distinct tissue types within the same individual. To investigate sXCI across tissues in the same individual, we used the GTEx dataset, which contains RNA-sequencing data for 4,571 individual tissues across 264 female donors. sXCI levels for each tissue were determined by measuring the allele-specific expression of reads arising from genes that undergo XCI, as recently described by us^6^. Whereas the average skewing across tissues per individual was highly similar to that observed at birth; with 12.89% of females highly skewed (median +1 SD, >71.4% skewing) for XCI and 6.25% exhibiting extreme skewing (median +2 SD, >79% skewing) (**Fig. 3a, b**), whole blood exhibited significantly higher levels of sXCI in older adults (**Fig. 3c**), consistent with the selective effects of clonal hematopoiesis. Notably, across all adults tested, constitutional skewing was the likely origin of 23.4% of extremely skewed females and in individuals under 50 years of age, constitutional skewing was still the dominant cause of extreme sXCI (**Fig. 3c**). Unexpectedly, sXCI was also highly prevalent across non-hematopoietic tissues, with 70% of females (181/256) having at least one extremely skewed tissue (**Fig. 3d**). In total, 13.6% (622/4571, ∼1 in 7) of tissues examined exhibited extreme sXCI with adrenal gland, whole blood, liver and pancreas among the most frequently skewed (**Fig. 3e**). Together, these results indicate that multi-tissue constitutive skewing is a common phenomenon in the normal human population.

**Figure 3.**
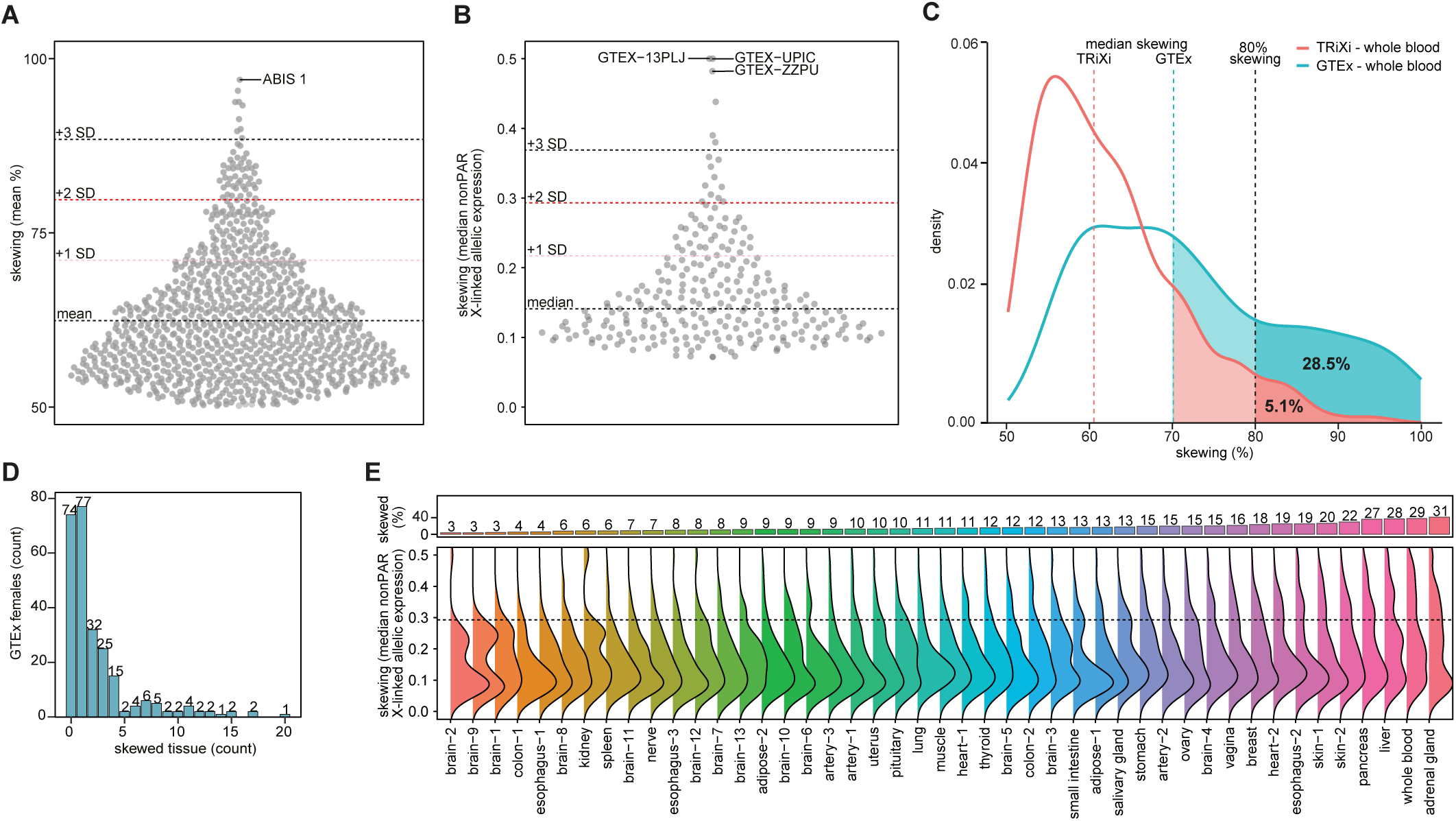
**Allelic-resolution analysis reveals widespread, multi-organ skewing in adults. a,b**, XCI status determined by TRiXi in 1,027 neonatal blood samples (a) and by median non-PAR X-linked allelic expression across 4,571 tissues in 264 female donors from the GTEx dataset (b). **c**, Density plot of skewing in whole blood as determined by TRiXi in newborns and allelic expression in adults from GTEx. **d**, bar plot showing adults grouped by the number of skewed tissues per individual. **e**, bar plot of fraction of extremely skewed (skewing >79%) samples per tissue (top) and ridge plot showing the distribution of skewing in all 45 tissues examined (bottom).

## DISCUSSION

We present TRiXi, a novel method for measurement of sXCI that offers considerable improvements over other techniques to detect sXCI, such as the HUMARA assay or next-generation sequencing approaches. TRiXi is highly sensitive (>99% sensitivity), accessible, affordable, and readily scalable for high-throughput use. Because the assay requires minimal DNA input and is compatible with short or degraded fragments, it is well-suited for screening samples with limited or compromised DNA, such as those commonly stored in biobanks. The *AR* locus was intentionally excluded from the TRiXi assay to avoid PCR bias associated with its broad normal repeat range. This also prevents unintended detection of premutation alleles linked to Kennedy’s disease, which are relatively common in the general population and often missed by the HUMARA assay due to preferential amplification of shorter alleles. Levels of sXCI assessed by TRiXi and HUMARA in the same samples were highly correlated, reinforcing the validity of historical HUMARA-based measurements and addressing persistent concerns about their accuracy^20, 22, 32, 33^. This comparison also underscored the key limitation of the HUMARA assay, with 24% (12/50) of samples being uninformative, representing a substantial waste of resources, including precious and limited patient-derived material. We applied TRiXi to over 1,000 archived umbilical cord blood samples from the ABIS birth cohort; making it the largest study of neonatal XCI status to date. Our findings demonstrate that high (>70%) and extreme (>80%) levels of XCI skewing are common at birth, occurring more frequently than previously recognized (15.55%), validating and expanding upon the results of earlier, smaller studies using the HUMARA assay^34^. Applying TRiXi to peripheral blood samples collected from the same ABIS donors at 14-16 years of age (*N* = 50) revealed sXCI to be highly variable throughout childhood with the exception of those females skewed at birth, which were more likely to remain skewed or proceed to extreme skewing, suggesting an ongoing selective process. Thus, even by 14 years of age, ∼10% of healthy females are already highly clonal in their peripheral blood and an additional 14% proceeding clearly proceeding towards clonality.

The origin of sXCI in any given individual is often unknown and can have a complex etiology. Skewed XCI can result either from variants that directly influence the XCI process on one of the two X-chromosomes such as mutations in the *XIST* promoter^25^, balanced X:autosome translocations (primary selection) or indirectly from variants that confer a growth advantage to specific cells or clones, such as those associated with clonal hematopoiesis (secondary selection)^25–27^. However, low-pass whole genome sequencing, targeted sequencing of established clonal hematopoiesis genes, Sanger sequencing of the *XIST* promoter, and ONT long-read sequencing did not identify any known genetic features previously linked to sXCI. Given that the drivers and molecular mechanisms establishing XCI remain incompletely defined, our investigation may not have captured genetic alterations in genes not yet known to be essential for XCI. Further, sXCI can originate stochastically, where, by chance, the small number of progenitor cells establishing a given tissue all independently inactivate the same parental X chromosome^12, 31^. The absence of identifiable genetic drivers of sXCI at birth may suggest a stochastic origin; however, this interpretation remains tentative without data on XCI patterns across other tissues from the same individual.

We assessed multi-organ skewing in humans leveraging the extensive genomic and transcriptomic data available for 264 post-mortem female donors in the GTEx database. Utilizing heterozygous genetic variants in X-linked genes, XCI status could be determined by measuring allele-specific expression levels in each tissue. As expected, we observed increased levels of sXCI in blood, consistent with progressive effects of clonal hematopoiesis over time. Whereas blood was the most frequently skewed tissue in adults, it surprisingly represented only 17% of all skewed tissues, with seven out of ten adult females skewed in at least one tissue.

Skewed X-inactivation is an intriguing and poorly understood genetic phenomenon in humans. However, it is also a genetic trait with a profound effect on the lives of thousands of carriers of X-linked disease, often resulting in the unpredictable manifestation of life-threatening symptoms of typically male X-linked disorders in females^35–38^. These X-linked diseases show variable symptoms correlated to the degree of XCI skewing with higher expression of the diseased allele resulting in more severe phenotypes^39, 40^. While most causative X-linked mutations can be detected by genetic testing, their heterozygous nature and possible inactivation during XCI does not currently allow for prediction of disease prognosis. A reliable estimation of sXCI in combination with genetic screening would provide more accurate clinical diagnoses to allow for individual approaches for treatment and interventions. TRiXi will allow for rapid, cost effective, and sensitive assessment of XCI status in carriers of X-linked diseases. Furthermore, TRiXi will facilitate the inclusion of sXCI measures in large-scale studies of genetic association in humans. Our results identify sXCI as a common and often multi-organ feature of humans, with potentially profound implications for our understanding of both the process of XCI, but also the potential impact of X-linked traits in females. Our findings also highlight the importance of studying multiple tissues when trying to understand X-linked process in humans. We further showed that extremely skewed XCI is likely to have a genetic origin, which remains to be identified. It should be considered that such a molecular basis for extreme sXCI is likely to be heritable even though previous studies did not find a heritable component to moderate levels of sXCI^12, 15^.

## METHODS

### Study population

Umbilical cord blood samples (n=1,027) were obtained from All Babies in Southeast Sweden (ABIS) cohort. ABIS is a prospective birth cohort study inviting all 21,700 children born between October 1997 and October 1999 in the southeast region of Sweden to participate^23, 24^. For longitudinal analysis of sXCI, paired whole blood samples collected at 1 year (when cord blood was not available; n= 30 1-year samples/n= 20 cord blood samples) and at 15 years of age were used. This study was approved by the Swedish Ethical Review Authority (#2022-06443-02).

### Automated DNA isolation

Genomic DNA from frozen umbilical cord blood samples was extracted using the QIAcube system with the QIAamp DNA Blood Mini QIAcube Kit (Qiagen, 51126) according to the manufacturer’s instructions. In short, blood samples were thawed at room temperature followed by incubation of 200 µl blood with 20 µl proteinase K and 200 µl lysis buffer for 10 minutes at 56°C. Lysed samples were mixed with 200 µl 99.9% ethanol and transferred to QIAamp spin columns. After two washing steps, DNA was eluted in 200 µl elution buffer.

### Manual DNA isolation

DNA from umbilical cord blood samples that were dried due to long-term storage and from capillary blood at 1 year of age were extracted manually using the QIAamp DNA Blood Mini Kit (Qiagen, 51106). Samples were thawed at room temperature, and a clump of dried blood (approximately 2 mm²) was transferred to a fresh microcentrifuge tube using pipette tips or scalpels, as needed. Each sample was then treated with 20 µL of proteinase K and 200 µL of lysis buffer (Qiagen, 939011), followed by thorough vortexing. The tubes were incubated at 56°C and shaken at 800 rpm for up to 6 hours. Buffer volume and incubation time were adjusted as necessary to ensure complete lysis. Once the blood was fully dissolved and lysed, DNA was purified using spin columns according to the manufacturer’s instructions, with elution performed in 100 µL of elution buffer.

### DNA quantification and quality assessment

DNA was quantified using a NanoDrop™ 2000/2000c Spectrophotometer (Thermo Scientific, ND-2000). For dried blood samples of questionable quality, DNA integrity was evaluated on agarose and only non-or lowly degraded DNA was included in this study.

### Identification of informative X-linked tandem repeats

To identify X-linked repeats that were (i) polymorphic, (ii) within 300 bp of a *Hpa*II digestion site (CCGG), and (iii) 50% methylated, the human genome reference (hg38) X-chromosome was screened for tri-and tetra-nucleotide repeats (containing one or two C or G combined) with at least six repeats in succession. To assess repeat length variability, we used whole genome sequencing (WGS data from the GTEx database from 50 females. Briefly, WGS CRAM files were downloaded from AnVIL using the Gen3 client. The files were sorted and converted into fastq files using Samtools^41^ (v1.9). Reads containing previously identified repeats of interest were extracted through detection of the ten bases upstream of each given repeat and repeat length was calculated for each individual read to identify polymorphic repeats. Repeats with a similar length distribution to the *AR* repeat were determined as having sufficient heterozygosity. To identify 50% methylated tandem repeat sites 300bp from either end of the repeat, pre-processed whole-genome bisulfite sequencing (WGBS) methylation score and coverage bigwig files from four females were downloaded from BLUEPRINT^42^ (CD4+ alpha-beta T cells; S009W451, P586, S007DD51, P582). Bigwigs were converted to Wig and then to bed file format with bigWigToWig^43^ and wig2bed^44^, respectively. CpG methylation scores were calculated by taking the mean methylation score per CpG site across all four samples. CpG sites not covered in all samples were removed. Sites with a methylation score between 0.34 and 0.66 were considered as 50% methylated and retained. Repeats lacking a 50% methylated CCGG sites within 300bp of either end of the repeat were excluded. Utilizing this approach we identified 15 repeats (see **Supplementary Table 1**) which were further tested in the lab. From these repeats, we utilized a total of five repeats (TRiXi 1-5) to reduce the chance of completely uninformative samples.

### *Hpa*II digestion

Genomic DNA (20-40 ng) was digested at 37°C for 60 minutes using 1× rCutsmart buffer and 1U of methylation-sensitive restriction enzyme *Hpa*II per sample, as per the manufacturer’s instructions (New England Biolabs, NEB-R0171M). The enzyme was inactivated at 80°C for 20 minutes after digestion. Additionally, for each run, a blank sample and a highly skewed sample were included as positive and negative controls, respectively.

### PCR amplification

After *Hpa*II digestion polymorphic tri- and tetra-nucleotide repeats from digested and undigested samples were amplified by PCR using a Phusion High-Fidelity Polymerase (New England Biolabs, NEB-M0530L). PCR amplification mix contained 1× GC PCR buffer, 8 nM dNTP mix, 3% v/v DMSO, and 0.4 U Phusion Polymerase. 10% v/v TRiXi primer mix was added to the master mix. PCR was performed using the entire volume of digested DNA (2 µl). For each sample 2 µl of undigested DNA (20 – 40 ng) were also PCR amplified as an undigested control. PCR amplification was performed using the following thermocycler protocol: One cycle of 30 seconds at 98°C; 30 cycles of 10 seconds at 98°C, followed by 30 seconds at 63°C, followed by 40 seconds at 72°C; one cycle of 5 minutes at 72°C.

### Capillary electrophoresis

For fragment analysis, 1 µl of the PCR reaction was transferred to a 96-well plate and diluted in 8 µl Hi-Di Formamide (Thermo Fisher, 4311320) and 0.2 µl GeneScan 600 LIZ sizig standard (Thermo Fisher, 4408399) mix. Sample plates were denatured at 95°C for 5 minutes, cooled on ice, and resolved using a 3500 Genetic Analyzer (Thermo Fisher).

### Calculating skewing and repeat length

Peak area, peak height and nucleotide size was utilized to calculate skewing and repeat length was obtained using GeneMapper5 and calculations were performed in R. Briefly, the .fsa files obtained from the Genetic Analyzer 3500 were loaded into GeneMapper v5 (Thermo Fisher). Peaks were called using microsatellite analysis (default setting with LIZ600+normalization as ladder). The resulting table with peak information was exported and loaded into R where skewing was calculated. First, the two highest peaks (separated by at least 2.5 bases) for each undigested sample were identified and matched to the closest sized peaks in the digested sample, obtaining two peaks for both undigested and digested sample. Samples where the peaks were not distinguishable from each other (*i.e.,* the peaks were not separated by more than 2.5 bases) in the undigested sample were deemed homozygous and the repeat was excluded from skewing calculations for that sample. Skewing was assessed individually for each repeat, and the mean of all informative repeats were used to determine degree of skewing. The skewing calculation itself was performed as previously described^45^:

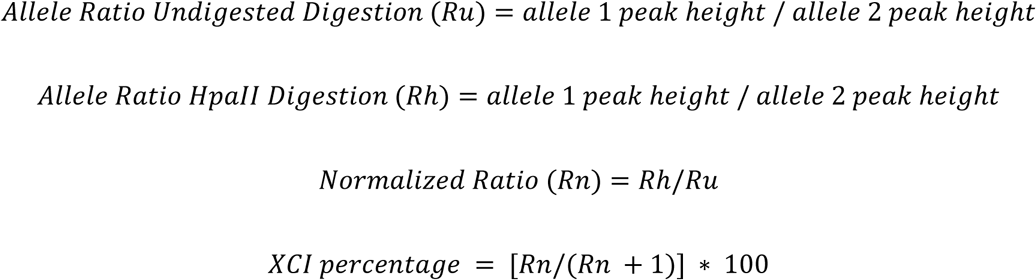

For each sample, the mean XCI percentage (ranging from 0 to 100%) was calculated, with 50% representing perfectly balanced XCI. XCI values were collapsed to a 50–100% range (as the direction of deviation from 50% is uninformative). Finally, the standard deviation and mean of the total data set was used to determine thresholds for skewing and extreme skewing:

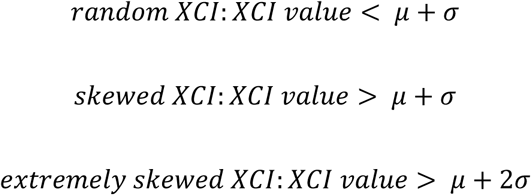

Where μ is the population mean and σ is the population standard deviation. Consequently, skewed XCI equated to values > 71.09% and extremely skewed XCI to values > 79.76% (**Fig. 2a**). XCI skew values above either threshold were classified as skewed, or extremely skewed, respectively. Finally, the repeat length was determined by comparing the obtained peak size to the expected PCR product length of a sample with a known number of repeats.

### Analysis of skewed X-inactivation in Genotype-Tissue Expression (GTEx) donors

Skewing was assessed from coupled RNA sequencing (RNA-seq) and whole exome sequencing data (WES) as previously described^6^. Briefly, WES BAM files for 256 female donors were downloaded from AnVIL using the Gen3 client. The files were sorted and converted into fastq files using Samtools (v1.9). Raw fastq reads were quality trimmed using FastP^46^ (v.0.20.0) with default settings. BWA-MEM^47^ was used for mapping reads to the human genome build 38 (hg38, GCA_000001405.15_GRCh38_no_alt_analysis_set.fna) using default settings. Sambamba^48^ v0.8.2 was used to mark duplicate reads. The GATK best practice germline short variant discovery pipeline was used to process the aligned reads, using base quality score recalibration and local realignment at known insertions and deletions (indels) (GATK v.4.2.6.1). Default filters were applied to indel and SNP calls using the variant quality score recalibration (VQSR) approach of GATK. All RNA-seq samples available for each participant (total of 4571 RNA-seq samples) were downloaded from AnVIL as aligned BAM files using the Gen3 client. Samtools (v1.9) was used to sort and convert the BAM files to fastq files and quality trimming was done with FastP (v.0.20.0) with default settings. RNA-seq reads were aligned with STAR^49^ 2-pass mode (v2.7.10a) with GCA_000001405.15_GRCh38_no_alt_analysis_set.fna and --sjdbGTFfile gencode.v40.annotation.gtf index. WASP filtering was performed to reduce reference bias^50^. Briefly, GATK SelectVariants was used to extract hetSNPs (that passed the VQSR filtering) for each individual from the WES VCFs and subsequently passed to STAR via --varVCFfile and–waspOutputMode. Reads failing WASP filtering were removed. Following data pre-processing, allele-specific expression analysis of the allele counts for hetSNPs was retrieved from RNA-seq data using GATK ASEReadCounter. Heterozygous variants that passed VQSR filtering were first extracted for each sample from WES VCFs using GATK SelectVariants. Sample-specific VCFs and RNA-seq BAMs were input to ASEReadCounter requiring minimum base quality of 20 in RNA-seq data. To remove potentially spurious sites, conservative filters were applied to the data (variant call read depth >= 10 per allele and minor allele read count > 10% of the total variant read count for WES data, and an RNA-seq read depth of at least seven reads). If a gene carried multiple hetSNPs, the hetSNP with the highest read count per tissue and individual was kept. Allele-specific expression was calculated as previously described^51^:

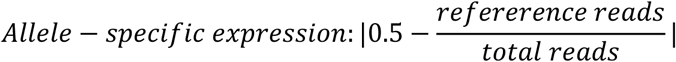

Skewing can then be calculated by taking the median allele-specific expression of all expressed non-pseudoautosomal (PAR) X-linked genes in each sample. The closer that value is to 0.5, the more preferential use of either parental allele. For instance, a female with a 100% skewed XCI would have a value of 0.5 while a female with a 50/50 usage of parental alleles would have 0. Similarly to how XCI skew was classified using TRiXi, the standard deviation and mean of the total data set was used to determine thresholds for skewing and extreme skewing:

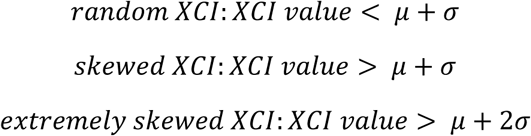

Consequently, skewed XCI equated to values >0.214 and extremely skewed XCI to values >0.29 (**Fig.2A**). XCI skew values above either threshold was classified as skewed, or extremely skewed, respectively.

### Low-pass whole genome sequencing

Whole genome DNA sequencing libraries were prepared with the NEBNext Ultra II DNA Library Prep Kit for Illumina (New England Biolabs, E7645S) and the NEBNext Multiplex Oligos for Illumina Index Primers Set 1 (New England Biolabs, E7335S). 200 ng of DNA were diluted to 50 µl in 1X TE (10 mM Tris-HCl (pH 8.0), 0.1 mM EDTA) and fragmented on the Bioruptor sonicator (Diagenode) to an average fragment size of 300 bp. End prep, sequencing adaptor ligation and PCR amplification with indexing primers were conducted according to the manufacturer’s instructions. Adaptor-ligated DNA was cleaned up and size selected for 250 – 400 bp fragments with AMPure XP beads (Beckman Coulter, A63881) by performing a two-step bead clean up with 25 µl beads in step one and subsequently adding 15 µl beads to the supernatant. Libraries were amplified for three PCR cycles. Quality and size of all libraries was assessed on the TapeStation using the D1000 ScreenTape (Agilent, 5067-5582) and corresponding reagents (Agilent, 5067-5583). Quantification was done with the 1X dsDNA High Sensitivity assay (Invitrogen, Q33231) on the Qubit fluorimeter. Libraries were sequenced on Illumina’s MiSeq machine using v2 reagents (Illumina, MS-102-2002) as 150bp paired-end reads. Each library was sequenced for a minimum of 2.4 million reads resulting in a coverage of >0.2x. Copy number analysis was performed using the R-package QDNASeq^52^. Briefly, the low pass WGS data was mapped to hg38 with BWA-MEM2^53^ using default parameters. Raw copy numbers were then estimated by counting the number of reads mapped to non-overlapping bins of 500kb.

### Sanger sequencing

Skewed ABIS samples were analyzed for mutations in the *XIST* minimal promoter region and the first exon by sanger sequencing. Briefly, 200 ng of sample DNA were amplified with FastStart Taq DNA Polymerase at 5 U/μl (Roche Applied Science, 12032902001). Appropriate DNA primers (Integrated DNA Technologies) were used at a concentration of 5 µM per reaction and designed to amplify the promoter region (Forward: 5’ TGG TTG GGGA GCC ATA CAA GG 3’, reverse: 5’ AGA ATG GGG GAA GGG ATA GG 3’) as well as the minimal promoter, including exon 1 (Forward: 5’ GGT GGA GGA GTT ACA AAT TCT GG 3’, reverse: 5’ TGC AGA GAG ATC TTC AGT CAG G 3’). The reaction mix was incubated on a SimpliAmp Thermal Cycler (Applied Biosystems, A24811) at 95°C for 4 minutes to allow enzyme activation. Thereafter, denaturation was performed at 95°C, followed by annealing at 62°C for 30 seconds each, and extension at 72°C for 60 seconds. This cycle profile was repeated 30 times before terminating with a final extension step for 7 minutes at 72°C. Amplified DNA was diluted to 50 ng/µL and cleaned using the DNA Clean & Concentrator-5 Kit (Zymo research, D4014) according to the manufacturers protocol. The DNA was washed twice and eluted in 20 µL of water. DNA concentration was determined using the NanoDrop™ 2000/2000c Spectrophotometer (Thermo Scientific, ND-2000) and adjusted to 5 ng per µL. Sanger Sequencing was performed at Eurofins genomics using the Mix2Seq LightRun service. The quality of the sequences obtained was inspected using Chromas 2.6.6 DNA sequencing software (Technelysium Pty Ltd, Australia). Sequence alignment was performed using the UCSC Genome browser’s BLAT tool as well as NCBI’s BLASTn to identify possible mutations.

### Identification of clonal hematopoiesis variants by Amplicon Sequencing

We performed amplicon sequencing with the help of the Pillar oncoReveal Myeloid Panel (Illumina, HDA-MY-1001-24), which includes 58 genes involved in clonal hematopoiesis^54^ following the protocol provided by the manufacturer (version 4.0). In brief, 40 ng of DNA was diluted in nuclease-free water to a total volume of 4.25 µl followed by targeted PCR amplification of coding sequences and mutation hotspot location of genes of interest using a target-specific pool of primers. Unused primers were removed by digestion with exonuclease I and clean-up of PCR products with AMPure XP magnetic beads (Beckman Coulter, A63881). Gene-specific amplicons were further PCR amplified for six cycles with Pillar Custom Indexing Primers Kit A, Indices PI501-6 and PI701-2 (Illumina, IDX-PI-1001-96). Libraries were quantified and quality controlled on the TapeStation with the D1000 ScreenTape and reagents (Agilent, 5067-5582 and 5067-5583). More than 2.4 million reads were sequenced per sample as 150 bp paired-end reads on the MiSeq sequencer (Illumina).

To identify variants associated with clonal hematopoiesis of indeterminate potential, we performed somatic variant calling in the amplicon sequencing data. Briefly, raw fastq reads were quality trimmed using fastp (0.24.0, default settings) and BWA-MEM2 was used to align reads to hg38 (GCA_000001405.15_GRCh38_no_alt_analysis_set.fna) using default settings. Sambamba v1.0.1 was used to mark duplicate reads. The GATK best practice ‘Somatic short variant discovery (SNVs + Indels)’ pipeline was used to process the aligned reads, using base quality score recalibration and local realignment at known insertions and deletions (indels) (GATK v.4.6.1.0). Default filters were applied to indel and SNP calls using the variant quality score recalibration (VQSR) approach of GATK. HaplotypeCaller, Mutect2, GetPileupSummaries, CalculateContamination and FilterMutectCalls (all default settings) were used to obtain analysis-ready vcf files with somatic variant calls. Only variants with ‘PASS’ filter status which had previously been reported as affecting clonal hematopoiesis^55, 56^ were considered to indicate clonal hematopoiesis.

### Oxford Nanopore long-read sequencing

1.5 µg of genomic DNA was sheared to approximately 10 kb using Covaris g-TUBEs (Covaris, 500291) in an Eppendorf 5415 R centrifuge at 5,000 RPM for 1 minute at room temperature. Library preparation was performed using the Ligation Sequencing Kit V14 (ONT, SQK-LSK114) according to the manufacturer’s protocol (GDE_9161_v114_revY, 30 January 2025). Briefly, DNA repair and end-preparation were conducted using the NEBNext FFPE DNA Repair Mix and Ultra™ II End Prep Enzyme Mix (NEB, E7672S) followed by adapter ligation. The final library was eluted and quantified using the Qubit 1X dsDNA High Sensitivity Assay (Thermo Fisher Scientific, Q33231). A total of 50 fmol of the prepared library was loaded onto a PromethION Flow Cell (ONT, FLO-PRO114) and sequenced on the PromethION 2 integrated device for 72 hours. The sequencing run generated 20.4 million reads at ∼46X coverage with a basecalled N50 read length of approximately 10kbp. The long-read sequencing was performed in collaboration with Clinical Genomics Linköping, SciLifeLab, who also provided technical support.

Determining XCI skew from the ONT long-read data was performed as previously described^17^. POD5 files were basecalled and aligned to the human genome build 38 (hg38, GCA_000001405.15_GRCh38_no_alt_analysis_set.fna) using dorado v0.7.4 (https://github.com/nanoporetech/dorado, which utilizes minimap2^57^ for mapping) and the dna_r10.4.1_e8.2_400bps_sup@v4.3.0 model. For modified basecalling, 5mC and 5hmC were called in a GC context. To allow phasing, high-quality variants were called X-linked variants were called using DeepVariant^58^ v1.8.0 using the model ‘ONT_R104’ and subsequent filtering with BCFtools^41^ (only ‘PASS’ variants were kept). Phasing of the VCF and subsequent haplotag assignment was performed using LongPhase^59^ v1.7.3. Reads assigned to incorrect haplotype blocks were identified in Integrative Genomics Viewer^60^ (v. 2.19.4) and subsequently corrected. Each finalized modbam file (with haplotags) was then processed individually in R using NanoMethViz^61^ v3.2.0. Methylation probabilities for each CCGG site in the TRiXi 1-5 PCR products were extracted per read, using the ‘query_methy’ function. Methylation probabilities were classified as low (and thus, associated with the active X (Xa) if below 0.1 and high (and associated with the Xi) if above 0.9. Within each repeat, the number of haplotype 1/Xa, haplotype 2/Xi, haplotype 1/Xi, and haplotype 2/Xa reads were counted to estimate the repeat-wise skew. To assess XCI skew, the mean of all repeat-wise skew values was used.

### Code Availability

Software for analysis of TRiXi data and the GTEx data is available on github: https://github.com/ColmNestorLab/skewing_XCI.

### Data availability

All sequencing data generated in this study have been deposited in the Swedish National Data Service (SND, https://snd.gu.se/, a data repository certified by Core Trust Seal). Access to restricted data will require completion of a Data Access Request (DAR) via the SND website. Each Data Access Request will be evaluated individually according to Swedish legislation. Source data are provided with this paper.

## Acknowledgements

We acknowledge the Core Facility at the Faculty of Medicine and Health Sciences, Linköping University, for their support with equipment and assistance for sequencing. This work was funded by the European Research Council (XX-Health-101045171).

## Author information Contributions

C.E.N. managed and designed the study. D.G., M.B., I.J., J.G., J.K., L.H., and M.V. performed experiments. D.G., B.G., M.B. and C.E.N analyzed the data. B.G. performed all bioinformatics analysis. B.G., E.H., S.J., S.H., S.L., M.B., J.L. and C.E.N wrote the manuscript and prepared the figures.

## Ethics declaration Competing interests

C.E.N has filed a patent application for the TRiXi method. The authors declare no other competing interests.

## SUPPLEMENTARY FIGURE LEGENDS

**Supplementary Figure 1.**
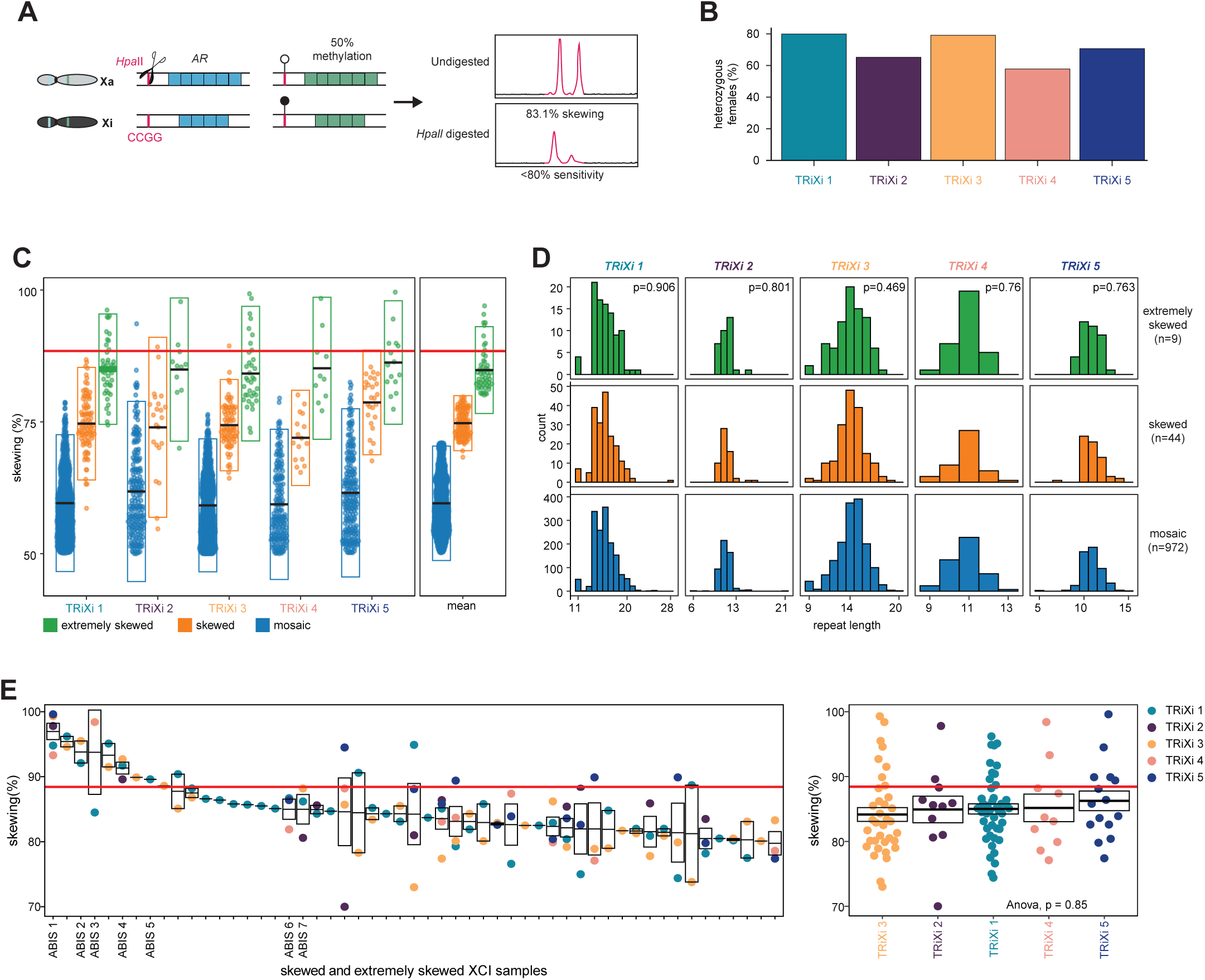
a, overview of the HUMARA assay based on the polymorphic CAG repeat in the *AR* gene. **b,** fraction of samples that were heterozygous for TRiXi repeat 1-5. **c**, boxplot of skewing of the five TriXi repeats across the three XCI skew groups. **d**, distribution of repeat lengths identified with TRiXi for repeat 1-5, split on XCI skew status. A Kolmogorov–Smirnov (K-S) test was used to statistically test if there was a difference between the XCI skew groups. **e**, skewing per individual (left) and per repeat (right), from samples classified as ‘skewed’ or ‘extremely skewed’ samples. ANOVA test; ρ = 0.85.

## SUPPLEMENTARY TABLES

Supplementary Table 1. Highly polymorphic tri- and tetra-nucleotide X-linked repeat sequences flanked by 50% methylated HpaII sites tested by multiplexing for suitability in TRiXi.

Supplementary Table 2. PCR primers used to amplify the 5 most suitable repeat sequences included in TRiXi.

Supplementary Table 3. Skewing of 50 ABIS-samples as measured using the AR-repeat based HUMARA assay, rendering 24% inconclusive samples due to homozygosity at the repeat.

Supplementary Table 4. TRiXi-based assessment of allelic skewing in 365 ABIS-samples, with conclusive results showing no individuals homozygous across all five repeat loci.

Supplementary Table 5. Skewing of ABIS-samples as measured by mini-TRiXi using 2 out of 5 repeats.

Supplementary Table 6. Longitudinal analysis of allelic skewing at age 0-1 year and 14-16 years as determined by the 2-repeat-based miniTRiXi for 50 ABIS-individuals.

Supplementary Table 7. Mutation analysis of genes associated with clonal hematopoiesis using the oncoRevealTM Myeloid Panel. 0= no mutation

